# Primary care practitioners’ preconception health literacy and information-seeking: A cross-sectional survey

**DOI:** 10.64898/2026.06.07.26355117

**Authors:** Cherie Caut, Danielle Schoenaker, Erica McIntyre, Amie Steel

**Author notes:** Corresponding author: Amie Steel.

## Abstract

**Background:** Parental health before pregnancy influences maternal and child outcomes. Primary care professionals, including general practitioners [GPs], midwives, and naturopaths, can provide preconception care, yet many report limited knowledge and difficulty accessing relevant information. This study described Australian GPs’, midwives’, and naturopaths’ preconception health literacy, including knowledge and ability to access information.

**Methods:** Between July and September 2022, Australian GPs, midwives, and naturopaths completed a 32-item online cross-sectional survey. Participants were recruited through professional associations, and data were analysed using descriptive and inferential statistics

**Results:** Participants (N=373) included naturopaths (40.7%), GPs (32.4%), and midwives (26.8%). Reported barriers to clinician health literacy including lack of preconception care resources (25.5%), and limited clinician knowledge (23.6%). The proportion identifying limited clinician knowledge differed significantly between professions (GP: 31.4%; midwives: 23.0%; naturopaths: 17.8%; p=0.030). The highest level of accurate knowledge regarding preconception exposures was for pre-pregnancy obesity (82.7%), while low birth weight was the most accurately identified preconception outcomes (83.7%). Incorrect responses were most common for maternal multivitamin use as an exposure (28.3%) and childhood leukaemia as an outcome (26.3%). Differences between professions were strongest for infant outcomes, with moderate associations observed for shoulder dystocia (V=.2355), precipitous labour (V=.2173), macrosomia (V=.2060), labour dystocia (V=.2018) and cryptorchidism (V=.2018).

**Discussion:** Preconception health literacy varies across primary care professions. Clinicians require greater access to targeted resources and education tailored to their differing scopes of practice and experience. Improving clinician preconception health literacy may strengthen consistent evidence-based care and support better maternal, child, and long-term family health outcomes.

## Introduction

Before pregnancy, parental health status influences health outcomes for both mother and child.^1–4^ Substantive evidence from systematic reviews describes multiple maternal and paternal modifiable preconception risks and health behaviours, including body composition, lifestyle behaviours, nutrition, environmental exposures and birth spacing. These modifiable preconception risks and health behaviours influence parental and child health outcomes.^1–3^ Primary care offers an important opportunity for preconception care, through risk screening and implementing intervention strategies to people of reproductive age to optimise health before conception.^5^ Preconception counselling and education delivered by primary care professionals are effective at reducing risks and encouraging health-promoting behaviours such as taking folic acid-containing multivitamin supplements, consuming a healthy diet, increasing physical activity and reducing the consumption of substances such as caffeine, smoking, alcohol and illicit drugs.^6^ As such, the timely provision of preconception care through primary care services can moderate risks and prevent adverse maternal and child health outcomes.^6–10^

A range of primary care professionals can and do provide preconception care.^6,11^ Among the diverse primary care professions, general practitioners (GPs) are well placed to provide preconception care by having contact with the general population and are considered the leading providers.^12–14^ Midwives have contact with women between pregnancies and are therefore well-placed to provide preconception care to women planning another pregnancy in the future.^13,15–17^ Women in Australia also seek preconception health support from naturopaths^11^ (primary care clinicians that treat diverse health conditions and populations using a range of non-pharmacological treatments, often as the main health provider^18^).

Barriers to delivering preconception care reported by primary care practitioners include, but are not limited to, insufficient knowledge about preconception health, and lack of awareness of preconception risks and clinical practice guidelines.^19–21^ Clinicians have continued to identify the need for further training on preconception health and access to information resources on preconception risks and interventions to improve their confidence in delivering preconception care.^21^ These gaps in knowledge and in the ability to access information pertaining to a particular health topic are defined as ‘health literacy’ which encompasses both knowledge of a health topic, and the skills that individuals can use to identify and implement knowledge.^22^ Health professionals’ preconception health literacy therefore refers to their current knowledge regarding preconception health risks and outcomes as well as their ability to find and use preconception health information to support their clinical practice.

While health professionals have continued to identify barriers to accessing preconception health information and uncertainty about their own preconception care knowledge, little research has examined the specific preconception health literacy needs of different primary care professions. Greater understanding of profession-specific knowledge gaps and information needs is necessary to support consistent, evidence-based preconception care. As such, this study aimed to explore Australian GPs’, midwives’, and naturopaths’ preconception information-seeking behaviour and preconception health literacy.

## Materials and methods

### Study design and setting

A quantitative cross-sectional survey was conducted through the Qualtrics platform between July and September 2022. The participants’ professional associations including the Royal Australian College of General Practitioners (RACGP) Specific Interest group Antenatal and Postnatal Care, Australian College of Midwives (ACM), Naturopaths and Herbalists Association of Australia (NHAA) and Complementary Medicine Association (CMA) shared the survey link to their members via electronic direct mail and social media channels.

### Participants

Participant eligibility criteria included practising in one of three health professions: general practice, midwifery or naturopathy. Participants must have been in clinical practice in their profession during the previous twelve months and be providing their health services in Australia. These health professionals were invited to share their knowledge of modifiable preconception risks and health behaviours, their perspectives and experiences regarding information-seeking for preconception health information, and their perceptions regarding the ideal delivery of preconception care services. The survey required fifteen to twenty minutes to be completed in full. An incentivisation prize was offered to participants to complete the study comprising three gift vouchers, one for each health profession group.^23,24^

### Survey Instrument

The 32-item survey (see Supplementary File 1. Survey Instrument) comprised eight domains of which four were used to inform this analysis: (1) participant characteristics; (2) preconception care information access and knowledge use, and barriers and enablers to delivering preconception care; (3) knowledge of preconception risks or exposures impacting health outcomes; (4) knowledge of health outcomes resulting from preconception risks or exposures. Items testing participant’s preconception knowledge (domains 3 and 4) were based on research included in systematic reviews of evidence ^1^. All items in these sections had documented evidence of association, so no false associations were presented. Individuals external to the research team and reflective of the target population pilot tested the survey instrument for face validity and technical functionality. Minor amendments were made to the survey flow and item structure based on the feedback from the pilot testers.

#### Participant characteristics

Items in this domain collected data regarding participants’ gender, age, current practice, clinical practice experience, clinical practice setting, and the populations to which they provide health services.

#### Preconception care information access and knowledge use, and barriers and enablers to delivering preconception care

Items in this domain collected data regarding the types of information and knowledge sources and knowledge resources used by participants to inform their preconception care practice. Items also asked participants to identify any barriers or enablers they experience to providing preconception care.

#### Knowledge of preconception risks or exposures influence outcomes

Items in this domain collected data on participants’ knowledge concerning preconception risks that may impact maternal, pregnancy, and offspring (fetal, infant, child) health outcomes. Responses for each list of preconception risks and associated health outcomes in each item were scaled on a 6-point Likert scale (*extremely unlikely, moderately unlikely, slightly unlikely, slightly likely, unsure; moderately likely and extremely likely*).

#### Knowledge of health outcomes from preconception risks or exposures

Items in this domain collected data on participants’ knowledge concerning specific pregnancy, post-natal, infant, and child outcomes associated with maternal and paternal preconception exposures. Responses for each list of preconception risks and associated health outcomes in each item were scaled on a 6-point Likert scale (*extremely unlikely, moderately unlikely, slightly unlikely, slightly likely, unsure; moderately likely and extremely likely*).

### Statistical methods

Survey response data were imported from Qualtrics into Stata 18 (StataCorp LLC) for cleaning, coding and analysis. Data resulting from 6-point Likert scales were collapsed into a 3-point scale (Unlikely = extremely unlikely, moderately unlikely, slightly unlikely; Unsure = unsure; Likely = slightly likely, moderately likely and extremely likely). Low health literacy was defined as indicating ‘unlikely’ or ‘unsure’ to survey items whereas as high health literacy was defined by ‘likely’ responses. Data were analysed using descriptive statistics (frequencies, percentages, mean and standard deviation) and inferential statistics for tests of association, including Pearson chi-square tests (comparing categorical variables) and analysis of variance (ANOVA) (comparing categorical and continuous variables). Statistical significance was set at p<0.05. The effect size of significant associations was determined using Cramer’s V. The effect size was classified as negligible association (.00 and under .10); weak association (.10 and under .20); moderate association (.20 and under .40); relatively strong association (.40 and under .60); strong association (.60 and under .80) and very strong association (.80 and under 1.00), as reported by Rea and Parker (2014).^25^

## Results

### Participant characteristics

The survey link was accessed by 422 health professionals, of which 373 completed survey items, achieving a participation rate of 88.4% for the study. Table 1 describes the participants’ characteristics. Three health professions were represented among the participants, comprising naturopaths (40.8%, n=152), GPs (32.4%, n=121), and midwives (27.1%, n=101). Most participants (37.8%) had five to ten years of clinical practice experience, followed by less than five years (35.4%). Most participants practised in clinics with other health professionals (54.4%) or in hospital settings (36.5%). Most participants identified as women (61.6%) and were 30 to 39 years old (37%) or 21 to 29 years old (35.9%). The most common population groups supported by health professionals were adult women (79.6%) followed by adult men (56.8%). Differences were identified between professions within each category of characteristic.

**Table 1.** Participant Characteristics.

| Participant Characteristics | All<br>(n=373)<br>n (%) | GP<br>(n=124)<br>n (%) | Midwives<br>(n=101)<br>n (%) | Naturopaths<br>(n=153)<br>n (%) | p |
| --- | --- | --- | --- | --- | --- |
| **Clinical Practice Experience (n=373)** |  |  |  |  |  |
| Less than 5 years | 132(35.4) | 31(25.6) | 42(42.0) | 59(38.8) | **<0.001** |
| 5 to 10 years | 141(37.8) | 62(51.2) | 42(42.0) | 37(24.3) |  |
| 11 to 15 years | 51(13.7) | 16(13.2) | 10(10.0) | 25(16.4) |  |
| More than 16 years | 49 (13.1) | 12 (9.9) | 6 (6.0) | 31 (20.4) |  |
| **Clinical practice setting (n=373)** |  |  |  |  |  |
| Clinical practice by myself | 110(29.5) | 16(9.1) | 21(21.0) | 73(48.0) | **<0.001** |
| Clinical practice with other HPs | 203(54.4) | 71(58.7) | 58(58.0) | 74(48.7) | 0.181 |
| Hospital setting | 136(36.5) | 58(48.0) | 52(52.0) | 26(17.1) | **<0.001** |
| Other | 2(0.5) | 1(0.8) | 0(0.0) | 1(0.6) | 0.680 |
| **Gender(n=372)** |  |  |  |  |  |
| Woman | 229(61.6) | 54(44.6) | 59(59.0) | 116(76.3) | **<0.001** |
| Man | 135(36.3) | 65(53.7) | 36(36.0) | 34(22.5) |  |
| Non-binary, gender diverse, or not disclosed | 8(2.1) | 2(1.7) | 5 (5.0) | 1(0.7) |  |
| **Age(n=373)** |  |  |  |  |  |
| 29 years or younger | 137 (36.7) | 48(39.7) | 53 (53.0) | 36 (23.7) | **<0.001** |
| 30 to 39 years | 139(37.0) | 52(43.0) | 39 (39.0) | 47 (30.9) |  |
| 40 to 49 years | 61(16.3) | 16(13.2) | 4 (4.0) | 41 (27.0) |  |
| 50 years or older | 37 (9.9) | 5 (4.1) | 4 (4.0) | 28 (18.4) |  |
| **Patient populations supported (n=373)\*** |  |  |  |  |  |
| adult women | 297(79.6) | 95(78.5) | 69(69.0) | 133(87.5) | **0.002** |
| adult men | 212(56.8) | 78(64.5) | 38(38.0) | 96(63.1) | **<0.001** |
| infants | 145(38.9) | 42(35.0) | 38(38.0) | 65(42.8) | 0.390 |
| children | 140(37.5) | 41(33.9) | 20(20.0) | 79(52.0) | **<0.001** |
| adolescents | 148(39.7) | 41(33.9) | 26(26.0) | 81(53.3) | **<0.001** |
| elderly | 90(24.1) | 24(19.8) | 13(13.0) | 53(35.0) | **<0.001** |
| marginalised populations | 53(14.2) | 16(13.2) | 8(8.0) | 29(19.1) | **0.045** |
| indigenous peoples | 62(16.6) | 17(14.0) | 13(13.0) | 32(21.0) | 0.159 |
| culturally- and linguistically-diverse (CALD) communities | 34(9.1) | 9(7.4) | 7(7.0) | 18(11.8) | 0.314 |
| other populations not listed | 8(2.1) | 2(1.6) | 1(1.0) | 5(3.3) | 0.425 |

\*participants could select more than one population group

### Preconception health information access and use

Participants reported preconception information access and use as described in Table 2. The most accessed information sources for preconception health information were professional journals for clinicians (65.7%) and clinical textbooks (n=211) (56.6%) with the latter being the only type of information source with a similar proportion of use across all health professions (p=0.412). GPs relied least on preconception information from their patients (15.0%), whereas midwives (17.0%) and naturopaths (36.2%) used medicinal product companies as a preconception health information source the least.

**Table 2.**
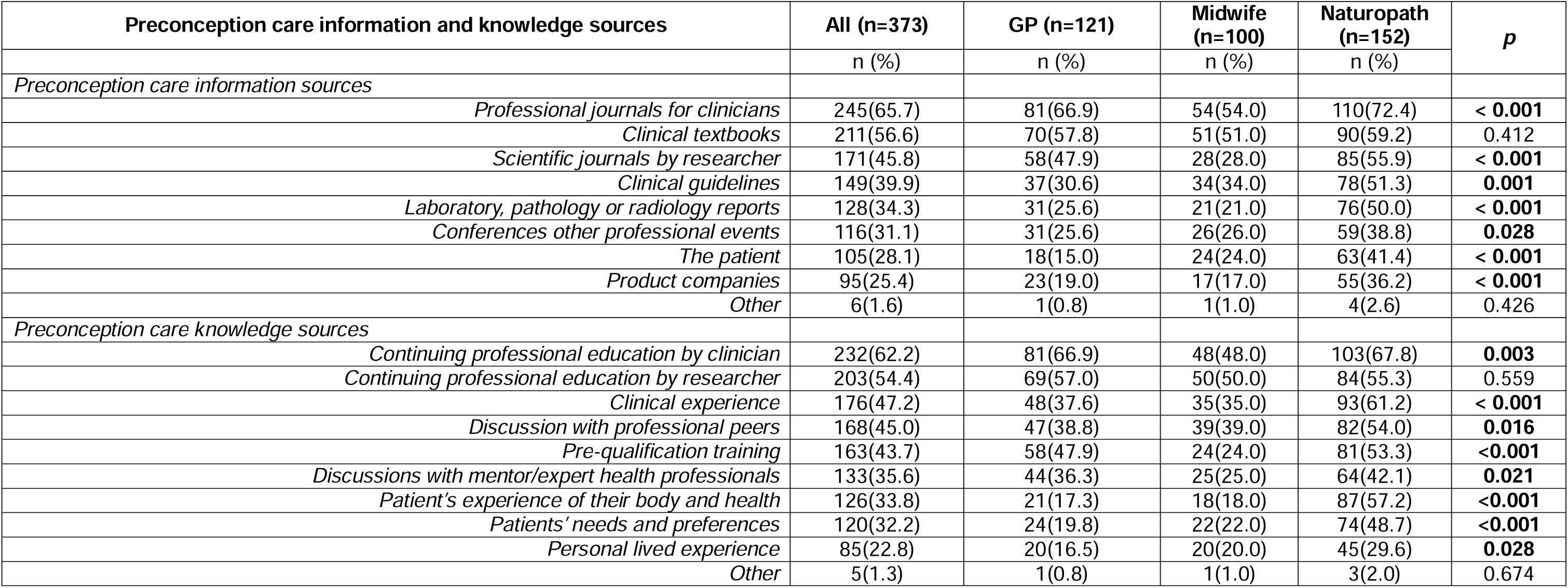
Preconception care information access and knowledge use.

| Preconception care information and knowledge sources | All (n=373) | GP (n=121) | Midwife (n=100) | Naturopath (n=152) | p |
| --- | --- | --- | --- | --- | --- |
|  | n (%) | n (%) | n (%) | n (%) |  |
| *Preconception care information sources* |  |  |  |  |  |
| *Professional journals for clinicians* | 245(65.7) | 81(66.9) | 54(54.0) | 110(72.4) | **< 0.001** |
| *Clinical textbooks* | 211(56.6) | 70(57.8) | 51(51.0) | 90(59.2) | 0.412 |
| *Scientific journals by researcher* | 171(45.8) | 58(47.9) | 28(28.0) | 85(55.9) | **< 0.001** |
| *Clinical guidelines* | 149(39.9) | 37(30.6) | 34(34.0) | 78(51.3) | **0.001** |
| *Laboratory, pathology or radiology reports* | 128(34.3) | 31(25.6) | 21(21.0) | 76(50.0) | **< 0.001** |
| *Conferences other professional events* | 116(31.1) | 31(25.6) | 26(26.0) | 59(38.8) | **0.028** |
| *The patient* | 105(28.1) | 18(15.0) | 24(24.0) | 63(41.4) | **< 0.001** |
| *Product companies* | 95(25.4) | 23(19.0) | 17(17.0) | 55(36.2) | **< 0.001** |
| *Other* | 6(1.6) | 1(0.8) | 1(1.0) | 4(2.6) | 0.426 |
| *Preconception care knowledge sources* |  |  |  |  |  |
| *Continuing professional education by clinician* | 232(62.2) | 81(66.9) | 48(48.0) | 103(67.8) | **0.003** |
| *Continuing professional education by researcher* | 203(54.4) | 69(57.0) | 50(50.0) | 84(55.3) | 0.559 |
| *Clinical experience* | 176(47.2) | 48(37.6) | 35(35.0) | 93(61.2) | **< 0.001** |
| *Discussion with professional peers* | 168(45.0) | 47(38.8) | 39(39.0) | 82(54.0) | **0.016** |
| *Pre-qualification training* | 163(43.7) | 58(47.9) | 24(24.0) | 81(53.3) | **<0.001** |
| *Discussions with mentor/expert health professionals* | 133(35.6) | 44(36.3) | 25(25.0) | 64(42.1) | **0.021** |
| *Patient's experience of their body and health* | 126(33.8) | 21(17.3) | 18(18.0) | 87(57.2) | **<0.001** |
| *Patients' needs and preferences* | 120(32.2) | 24(19.8) | 22(22.0) | 74(48.7) | **<0.001** |
| *Personal lived experience* | 85(22.8) | 20(16.5) | 20(20.0) | 45(29.6) | **0.028** |
| *Other* | 5(1.3) | 1(0.8) | 1(1.0) | 3(2.0) | 0.674 |

Participants’ preconception health knowledge was most often acquired through continuing professional education (CPE) provided by a clinician (62.2%) and researcher (54.4%). This researcher-delivered CPE training was the only specific preconception health knowledge source which each group of health professions had no difference in the proportion of use (p=0.559). While GPs and midwives most commonly reported acquiring preconception health knowledge through CPE provided by a clinician (GP: 66.9%; midwives: 48.0%) or researcher (GP: 57.0 %; midwives: 50.0%), naturopaths most often reported CPE provided by a clinician (67.8%), followed by their own clinical experience (61.2%). Preconception health knowledge was least often acquired through participants’ personal lived experience (22.8%) across all health professions, although the proportion of respondents for each group that identified this source was significantly different (GPs: 16.5%; naturopaths: 29.6%; midwives: 18.0%; p=0.028).

### Barriers to patient and clinician preconception health literacy

Participants reported insufficient patient preconception health literacy as seen through poor community awareness of preconception health in general (54.4%) and of the importance of preconception health (40.2%) (see Table 3). Less commonly, participant reported barriers associated with clinician health literacy including lack of clinical preconception care resources (25.5%), and of clinician preconception health knowledge (23.6%), alongside conflicting views of preconception care priorities among health professionals (23.6%). The lack of preconception care resources was consistent across all professions (p=0.535), but there was a significant difference in the proportion of participants identifying lack of clinician preconception health knowledge (GP: 31.4%; midwives: 23.0%; naturopaths: 17.8%; p=0.030), and those indicating conflicting health professionals’ views regarding preconception care priorities (GP: 19.8%; midwives: 15.0%; naturopaths: 32.2%; p=0.003) as barriers.

**Table 3:**
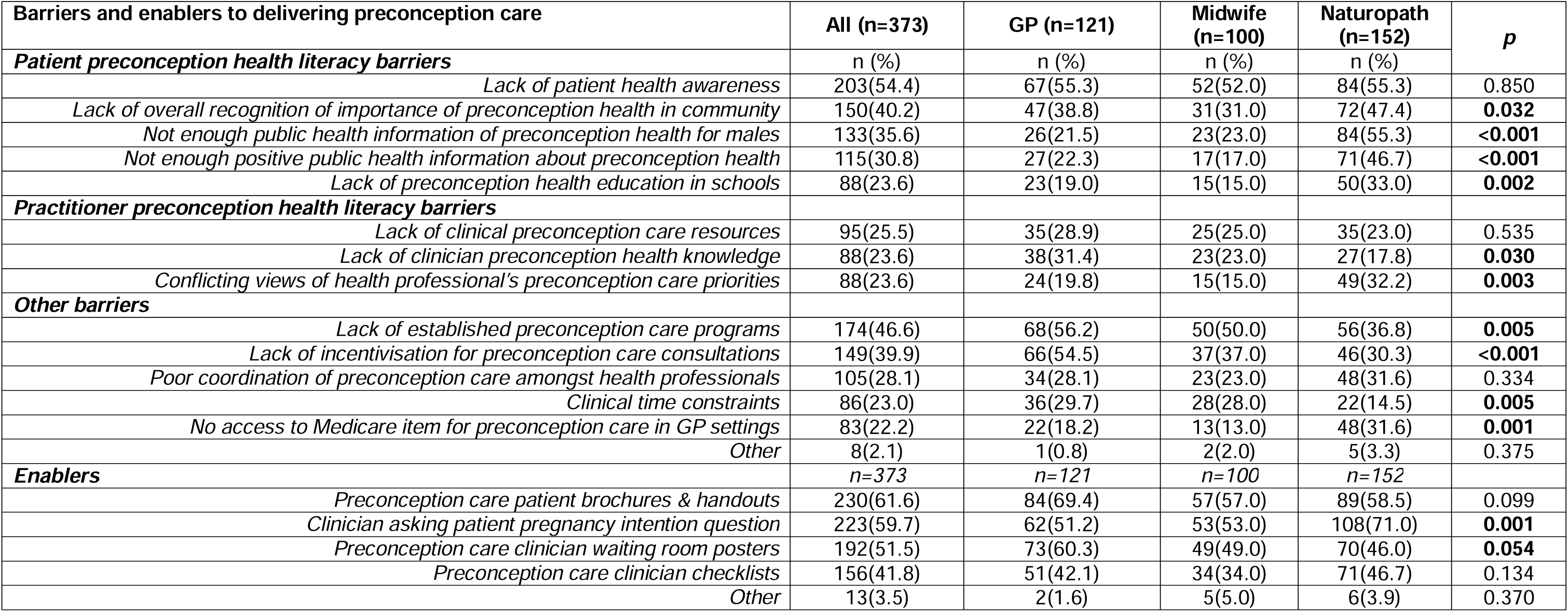
Barriers and enablers to delivering preconception care and preconception health literacy.

| Barriers and enablers to delivering preconception care | All (n=373) | GP (n=121) | Midwife (n=100) | Naturopath (n=152) | p |
| --- | --- | --- | --- | --- | --- |
| **Patient preconception health literacy barriers** | n (%) | n (%) | n (%) | n (%) |  |
| *Lack of patient health awareness* | 203(54.4) | 67(55.3) | 52(52.0) | 84(55.3) | 0.850 |
| *Lack of overall recognition of importance of preconception health in community* | 150(40.2) | 47(38.8) | 31(31.0) | 72(47.4) | **0.032** |
| *Not enough public health information of preconception health for males* | 133(35.6) | 26(21.5) | 23(23.0) | 84(55.3) | **<0.001** |
| *Not enough positive public health information about preconception health* | 115(30.8) | 27(22.3) | 17(17.0) | 71(46.7) | **<0.001** |
| *Lack of preconception health education in schools* | 88(23.6) | 23(19.0) | 15(15.0) | 50(33.0) | **0.002** |
| **Practitioner preconception health literacy barriers** |  |  |  |  |  |
| *Lack of clinical preconception care resources* | 95(25.5) | 35(28.9) | 25(25.0) | 35(23.0) | 0.535 |
| *Lack of clinician preconception health knowledge* | 88(23.6) | 38(31.4) | 23(23.0) | 27(17.8) | **0.030** |
| *Conflicting views of health professional's preconception care priorities* | 88(23.6) | 24(19.8) | 15(15.0) | 49(32.2) | **0.003** |
| **Other barriers** |  |  |  |  |  |
| *Lack of established preconception care programs* | 174(46.6) | 68(56.2) | 50(50.0) | 56(36.8) | **0.005** |
| *Lack of incentivisation for preconception care consultations* | 149(39.9) | 66(54.5) | 37(37.0) | 46(30.3) | **<0.001** |
| *Poor coordination of preconception care amongst health professionals* | 105(28.1) | 34(28.1) | 23(23.0) | 48(31.6) | 0.334 |
| *Clinical time constraints* | 86(23.0) | 36(29.7) | 28(28.0) | 22(14.5) | **0.005** |
| *No access to Medicare item for preconception care in GP settings* | 83(22.2) | 22(18.2) | 13(13.0) | 48(31.6) | **0.001** |
| *Other* | 8(2.1) | 1(0.8) | 2(2.0) | 5(3.3) | 0.375 |
| **Enablers** | n=373 | n=121 | n=100 | n=152 |  |
| *Preconception care patient brochures & handouts* | 230(61.6) | 84(69.4) | 57(57.0) | 89(58.5) | 0.099 |
| *Clinician asking patient pregnancy intention question* | 223(59.7) | 62(51.2) | 53(53.0) | 108(71.0) | **0.001** |
| *Preconception care clinician waiting room posters* | 192(51.5) | 73(60.3) | 49(49.0) | 70(46.0) | **0.054** |
| *Preconception care clinician checklists* | 156(41.8) | 51(42.1) | 34(34.0) | 71(46.7) | 0.134 |
| *Other* | 13(3.5) | 2(1.6) | 5(5.0) | 6(3.9) | 0.370 |

Participants perceived preconception care patient brochures and handouts would most enable preconception care (n=230, 61.6%), followed by clinicians asking patients about pregnancy intention (59.7%). A significant difference in the rate that clinicians asking the patient a question about their pregnancy intention was identified as enabling preconception care (GP: 51.2%; midwives: 53.0%; naturopaths: 71.0%; p=0.001).

### Knowledge of health outcomes from maternal preconception risks or exposures

Table 4 describes the participants’ reported knowledge of preconception risks and exposures. The highest level of accurate knowledge (i.e., ‘likely’) across the sample regarding preconception risks that influence maternal and pregnancy outcomes was for pre-pregnancy obesity (82.7%), smoking (82.3%), and poor diet (81.5%). The highest proportion of incorrect (i.e. ‘unlikely’) responses related to preconception risks impacting maternal and pregnancy outcomes were for maternal multivitamin use (28.3%), folic acid intake (27.1%) and lack of physical activity (18.6%). Participants responses demonstrating the highest level of uncertainty (i.e. ‘unsure’) for short inter-pregnancy interval impacting maternal and pregnancy outcomes (5.3%).

**Table 4.** Knowledge of the influence of maternal preconception exposures on health outcomes.

| Influence of maternal preconception exposures on... | Total |  |  | GPs |  |  | Midwives |  |  | Naturopaths |  |  | p value* | Cramer's V** |
| --- | --- | --- | --- | --- | --- | --- | --- | --- | --- | --- | --- | --- | --- | --- |
|  | Unlikely<br>n (%) | Unsure<br>n (%) | Likely<br>n (%) | Unlikely<br>n (%) | Unsure<br>n (%) | Likely<br>n (%) | Unlikely<br>n (%) | Unsure<br>n (%) | Likely<br>n (%) | Unlikely<br>n (%) | Unsure<br>n (%) | Likely<br>n (%) |  |  |
| **...maternal & pregnancy outcomes** |  |  |  |  |  |  |  |  |  |  |  |  |  |  |
| pre-pregnancy obesity (n=335) | 49<br>(14.6) | 9 (2.7) | 277 (82.7) | 17<br>(14.8) | 4 (3.5) | 94 (81.7) | 23<br>(24.2) | 4 (4.2) | 68<br>(71.6) | 9 (7.2) | 1 (0.8) | 115 (92.0) | .003 | .1553 |
| smoking (n=338) | 51<br>(15.1) | 9 (2.7) | 278 (82.3) | 21<br>(18.0) | 3 (2.6) | 93 (79.5) | 19<br>(20.2) | 5 (5.3) | 70<br>(74.5) | 11 (8.7) | 1 (0.8) | 115 (90.6) | .02 | .1320 |
| poor diet (n=340) | 56<br>(16.5) | 7 (2.1) | 277 (81.5) | 20<br>(17.0) | 3 (2.5) | 95 (80.5) | 26<br>(27.5) | 3 (3.2) | 66<br>(69.5) | 10 (7.9) | 1 (0.8) | 116 (91.3) | .002 | .1603 |
| pre-pregnancy overweight (n=338) | 55<br>(16.3) | 8 (2.4) | 275 (81.4) | 20<br>(17.0) | 4 (3.4) | 94 (79.7) | 20<br>(21.3) | 4 (4.3) | 70<br>(74.5) | 15 (11.9) | 0 (0.0) | 111 (88.1) | .06 |  |
| alcohol use (n=338) | 53<br>(15.7) | 11 (3.3) | 274 (81.1) | 23<br>(19.5) | 3 (2.5) | 92 (78.0) | 16<br>(17.2) | 6 (6.5) | 71<br>(76.3) | 14 (11.0) | 2 (1.6) | 111 (87.4) | .08 |  |
| exposure to ambient air pollution (n=337) | 55<br>(16.3) | 10 (3.0) | 272 (80.7) | 21<br>(17.8) | 4 (3.4) | 93 (78.8) | 16<br>(17.2) | 5 (5.4) | 72<br>(77.4) | 18<br>(14.3) | 1 (0.8) | 107 (84.9) | .301 |  |
| high caffeine intake (n=340) | 54<br>(15.9) | 12 (3.5) | 274 (80.6) | 21<br>(17.8) | 4 (4.4) | 93 (78.8) | 18<br>(19.0) | 5 (5.3) | 72<br>(75.8) | 15 (11.8) | 3 (2.4) | 109 (85.8) | .378 |  |
| lack of physical activity (n=339) | 63<br>(18.6) | 8 (2.4) | 268 (79.1) | 28<br>(23.7) | 4 (3.4) | 86 (72.9) | 19<br>(20.0) | 3 (3.2) | 73<br>(76.8) | 16<br>(12.7) | 1 (0.8) | 109 (86.5) | .102 |  |
| short inter-pregnancy interval (n=338) | 54<br>(16.0) | 18 (5.3) | 266 (78.7) | 19<br>(16.1) | 4 (3.4) | 95 (80.5) | 18<br>(19.0) | 5 (5.3) | 72<br>(75.8) | 17<br>(13.6) | 9 (7.2) | 99 (79.2) | .595 |  |
| pre-pregnancy underweight (n=338) | 63<br>(18.6) | 10 (3.0) | 265 (78.4) | 23<br>(19.7) | 3 (2.6) | 91 (77.8) | 26<br>(27.4) | 4 (4.2) | 65<br>(68.4) | 14 (11.1) | 3 (2.4) | 109 (86.5) | .03 | .1261 |
| folic acid intake (n=339) | 92<br>(27.1) | 9 (2.7) | 238 (70.2) | 18<br>(15.4) | 4 (3.4) | 95 (81.2) | 25<br>(26.3) | 4 (4.2) | 66<br>(69.5) | 49<br>(38.6) | 1 (0.8) | 77 (60.6) | .0001 | .1655 |
| Multivitamin intake (n=339) | 96<br>(28.3) | 11 (3.2) | 232 (68.4) | 21<br>(17.8) | 5 (4.2) | 92 (78.0) | 30<br>(31.6) | 4 (4.2) | 61<br>(64.2) | 45<br>(35.7) | 2 (1.6) | 79 (62.7) | .02 | .1304 |
| **...on offspring outcomes** |  |  |  |  |  |  |  |  |  |  |  |  |  |  |
| alcohol use (n=337) | 51<br>(15.1) | 10 (3.0) | 110 (87.3) | 21<br>(18.0) | 4 (3.4) | 92 (78.6) | 15<br>(16.0) | 5 (5.3) | 74<br>(78.7) | 15 (11.9) | 1 (0.8) | 110 (87.3) | .193 |  |
| exposure to ambient air pollution (n=336) | 52<br>(15.5) | 8 (2.4) | 276 (82.1) | 20<br>(17.1) | 4 (3.4) | 93 (79.5) | 17<br>(18.3) | 4 (4.3) | 72<br>(77.4) | 15 (11.9) | 0 (0.0) | 111 (88.1) | .108 |  |
| high caffeine intake (n=336) | 52<br>(15.5) | 10 (3.0) | 274 (81.6) | 21<br>(18.1) | 3 (2.6) | 92 (79.3) | 18<br>(19.4) | 6 (6.5) | 69<br>(74.2) | 13<br>(10.2) | 1 (0.8) | 113 (90.0) | .025 | .1285 |
| poor diet (n=337) | 58<br>(17.2) | 6 (1.8) | 273 (81.0) | 23<br>(19.7) | 3 (2.6) | 91 (77.8) | 22<br>(23.4) | 3 (3.2) | 69<br>(73.4) | 13<br>(10.3) | 0 (0.0) | 113 (90.0) | .02 | .1316 |
| smoking (n=336) | 55<br>(16.4) | 11 (3.3) | 270 (80.4) | 25<br>(21.4) | 4 (3.4) | 88 (75.2) | 18<br>(19.4) | 5 (5.4) | 70<br>(75.3) | 12 (9.5) | 2 (1.6) | 112 (88.9) | .039 | .1225 |
| pre-pregnancy obesity (n=335) | 60<br>(17.9) | 7 (2.1) | 268 (80.0) | 23<br>(19.8) | 3 (2.6) | 90 (77.6) | 23<br>(24.5) | 3 (3.2) | 68<br>(72.3) | 14 (11.2) | 1 (0.8) | 110 (88.0) | .059 |  |
| pre-pregnancy underweight (n=337) | 63<br>(18.7) | 11 (3.3) | 263 (78.0) | 22<br>(19.0) | 5 (4.3) | 89 (76.7) | 21<br>(22.3) | 4 (4.3) | 69<br>(73.4) | 20<br>(15.8) | 2 (1.6) | 105 (82.7) | .451 |  |
| pre-pregnancy overweight (n=336) | 73<br>(21.7) | 9 (2.7) | 254 (75.6) | 26<br>(22.4) | 3 (2.6) | 87 (75.0) | 27<br>(28.7) | 4 (4.3) | 63<br>(67.0) | 20<br>(15.9) | 2 (1.6) | 104 (82.5) | .121 |  |
| short inter-pregnancy interval (n=337) | 68<br>(20.2) | 18 (5.3) | 251 (74.5) | 27<br>(23.1) | 4 (3.4) | 86 (73.5) | 22<br>(23.7) | 7 (7.5) | 64<br>(68.8) | 19<br>(15.0) | 7 (5.5) | 101 (79.5) | .262 |  |
| lack of physical activity (n=337) | 78<br>(23.2) | 9 (2.7) | 250 (74.2) | 31<br>(26.5) | 4 (3.4) | 82 (70.1) | 25<br>(26.6) | 4 (4.3) | 65<br>(69.2) | 22<br>(17.5) | 1 (0.8) | 103 (81.8) | .128 |  |

|  | Total |  |  | GPs |  |  | Midwives |  |  | Naturopaths |  |  | *p* | *Cramer's* |
| --- | --- | --- | --- | --- | --- | --- | --- | --- | --- | --- | --- | --- | --- | --- |
| *folic acid intake (n=338)* | 93<br>(27.5) | 8 (2.4) | 237 (70.1) | 22<br>(18.8) | 4 (3.4) | 91 (77.8) | 22<br>(23.4) | 4 (4.3) | 68<br>(72.3) | 49<br>(38.6) | 0 (0.0) | 78 (61.4) | **.002** | .1579 |
| *multivitamin intake (n=333)* | 93<br>(27.9) | 10 (3.0) | 230 (69.1) | 19<br>(16.5) | 4 (3.5) | 92 (80.0) | 29<br>(30.9) | 4 (4.3) | 61<br>(64.9) | 45<br>(36.3) | 2 (1.6) | 77 (62.1) | **.01** | .1409 |

\*Unlikely = Extremely unlikely and moderately unlikely; Unsure = Slightly unlikely, slightly likely and unsure; Likely = Moderately likely and extremely likely
\*\*The Cramer's V and P values reflect statistical tests examining differences in health literacy between health professions

The analysis of all participants’ responses concerning the influence of maternal preconception exposures on offspring (fetal, infant or child) health outcomes found the highest level of knowledge was for maternal alcohol use (87.3%), exposure to ambient pollution (82.1%), high caffeine intake (81.6%), and poor diet (81.0%). The highest proportion of incorrect (‘unlikely’) responses were for maternal multivitamin (27.9%) or folic acid (27.5%) intake while the responses demonstrating the highest level of uncertainty were associations between offspring health outcomes short inter-pregnancy interval (5.3%).

Weak associations were identified between health profession group and accuracy of identified preconception exposures that impact maternal and pregnancy or offspring outcomes for several maternal exposures. The highest effect size for both outcome categories was identified for folic acid intake (maternal and pregnancy: V=.1655; offspring: V=.1579), followed by poor diet (maternal and pregnancy: V=.1603; offspring: V=.1316). GPs more commonly indicated folic acid as an exposure impacting both populations (maternal and pregnancy: GP 81.2%, midwives 69.5%, naturopaths 60.5%; offspring: GP 77.8%, midwives 72.3%, naturopaths 61.4%). Naturopaths most commonly identified poor diet as likely to impact outcomes for both populations (maternal and pregnancy: GP 80.5%, midwives 69.5%, naturopaths 91.3%; offspring: GP 77.8%, midwives 73.4%, naturopaths 90.0%).

### Knowledge of health outcomes from maternal preconception risks or exposure

The analysis of all participants’ responses concerning the influence of maternal preconception risks on pregnancy and post-natal outcomes (see Table 5) found the highest level of accurate knowledge was for antenatal depression (82.4%), reduced embryonic development (82.4%), and obesity (82.1%) as outcomes likely to result from maternal preconception exposures. The pregnancy and postnatal outcomes attributed with the highest level of uncertainty (‘unsure’) from maternal exposures were uterine rupture (6.4%) and placental abruption (6.1%). The greatest proportion of participants identified gestational diabetes as an unlikely outcome from maternal preconception exposures (18.0%).

**Table 5:** Knowledge of health outcomes from maternal preconception risks or exposures.

| Influence of maternal preconception exposures on... | Total |  |  | GP |  |  | Midwives |  |  | Naturopaths |  |  | p** | Cramer's V** |
| --- | --- | --- | --- | --- | --- | --- | --- | --- | --- | --- | --- | --- | --- | --- |
|  | Unlikely<br>n (%) | Unsure<br>n (%) | Likely<br>n (%) | Unlikely<br>n (%) | Unsure<br>n (%) | Likely<br>n (%) | Unlikely<br>n (%) | Unsure<br>n (%) | Likely<br>n (%) | Unlikely<br>n (%) | Unsure<br>n (%) | Likely<br>n (%) |  |  |
| **...pregnancy and post-natal outcomes** |  |  |  |  |  |  |  |  |  |  |  |  |  |  |
| antenatal depression (n=330) | 47<br>(14.2) | 11 (3.3) | 272 (82.4) | 17<br>(14.8) | 3 (2.6) | 95 (82.6) | 19<br>(20.2) | 3 (3.2) | 72 (76.6) | 11 (9.1) | 5 (4.1) | 105 (86.8) | .221 |  |
| reduced embryonic development (n=324) | 47<br>(14.5) | 10 (3.1) | 267 (82.4) | 19<br>(16.7) | 3 (2.6) | 92 (80.7) | 12<br>(13.3) | 3 (3.3) | 75 (83.3) | 16<br>(13.3) | 4 (3.3) | 100 (83.3) | .946 |  |
| obesity (n=329) | 52<br>(15.8) | 7 (2.1) | 270 (82.1) | 14<br>(12.3) | 1 (0.9) | 99 (86.8) | 21<br>(22.3) | 3 (3.2) | 70 (74.5) | 17<br>(14.1) | 3 (2.5) | 101 (83.5) | .193 |  |
| postnatal depression (n=330) | 54<br>(16.4) | 10 (3.0) | 266 (80.6) | 22<br>(19.1) | 3 (2.6) | 90 (78.2) | 18<br>(19.2) | 3 (3.2) | 73 (77.7) | 14 (11.6) | 4 (3.3) | 103 (85.1) | .513 |  |
| preeclampsia (n=328) | 56<br>(17.1) | 8 (2.4) | 264 (80.5) | 21<br>(18.3) | 2 (1.7) | 92 (80.0) | 18<br>(19.4) | 4 (4.3) | 71 (76.3) | 17<br>(14.2) | 2 (1.7) | 101 (84.2) | .521 |  |
| pregnancy induced hypertension (n=329) | 53<br>(16.1) | 12 (3.7) | 264 (80.2) | 19<br>(16.7) | 2 (1.8) | 93 (81.6) | 18<br>(19.2) | 5 (5.3) | 71 (75.5) | 16<br>(13.2) | 5 (4.1) | 100 (82.6) | .481 |  |
| gestational diabetes (n=328) | 59<br>(18.0) | 7 (2.1) | 262 (80.0) | 26<br>(22.6) | 1 (0.9) | 88 (76.5) | 21<br>(22.3) | 3 (3.2) | 70 (74.5) | 12<br>(10.1) | 3 (2.5) | 104 (87.4) | .055 |  |
| miscarriage (n=330) | 54<br>(16.4) | 13 (3.9) | 263 (79.7) | 19<br>(16.5) | 2 (1.7) | 94 (81.7) | 17<br>(18.1) | 7 (7.5) | 70 (74.5) | 18<br>(14.9) | 4 (3.3) | 99 (81.8) | .262 |  |
| uterine rupture (n=329) | 58<br>(17.6) | 21 (6.4) | 250 (76.0) | 18<br>(15.7) | 3 (2.6) | 94 (81.7) | 20<br>(21.3) | 6 (6.4) | 68 (72.3) | 20<br>(16.7) | 12 (10.0) | 88 (73.3) | .145 |  |
| placental abruption (n=328) | 63<br>(19.2) | 20 (6.1) | 245 (74.7) | 24<br>(21.1) | 2 (1.8) | 88 (77.2) | 18<br>(19.4) | 5 (5.4) | 70 (75.3) | 21<br>(17.4) | 13 (10.7) | 87 (71.9) | .074 |  |
| **...infant outcomes** |  |  |  |  |  |  |  |  |  |  |  |  |  |  |
| low birth weight (n=331) | 48<br>(14.5) | 6 (1.8) | 277 (83.7) | 21<br>(18.3) | 0 (0.0) | 94 (81.7) | 16<br>(17.0) | 4 (4.3) | 74 (78.7) | 11 (9.0) | 2 (1.6) | 109 (89.3) | .039 | .1233 |
| small for gestational age (n=328) | 57<br>(17.4) | 5 (1.5) | 266 (81.1) | 19<br>(17.0) | 0 (0.0) | 93 (83.1) | 22<br>(23.4) | 3 (3.2) | 69 (73.4) | 16<br>(13.1) | 2 (1.6) | 104 (85.3) | .104 |  |
| neural tube defects (n=330) | 56<br>(17.0) | 8 (2.4) | 266 (80.6) | 22<br>(19.1) | 0 (0.0) | 93 (80.9) | 20<br>(21.5) | 4 (4.3) | 69 (74.2) | 14 (11.5) | 4 (3.3) | 104 (85.3) | .063 |  |
| preterm birth (n=327) | 63<br>(19.3) | 7 (2.1) | 257 (78.6) | 21<br>(18.4) | 0 (0.0) | 93 (81.6) | 27<br>(29.0) | 3 (3.2) | 63 (67.7) | 15<br>(12.5) | 4 (3.3) | 101 (84.2) | .01 | .1423 |
| large for gestational age (n=330) | 61<br>(18.5) | 10 (3.0) | 259 (78.5) | 20<br>(17.5) | 0 (0.0) | 94 (82.5) | 17<br>(18.1) | 4 (4.3) | 73 (77.7) | 24<br>(19.7) | 6 (4.9) | 92 (75.4) | .207 |  |
| other congenital abnormalities (n=329) | 63<br>(19.2) | 11 (3.3) | 255 (77.5) | 23<br>(20.0) | 0 (0.0) | 92 (80.0) | 23<br>(24.7) | 4 (4.3) | 66 (71.0) | 17<br>(14.1) | 7 (5.8) | 97 (80.2) | .038 | .1243 |
| admission to neonatal intensive care (n=330) | 60<br>(18.2) | 17 (5.2) | 253 (76.7) | 21<br>(18.3) | 0 (0.0) | 94 (81.7) | 19<br>(20.4) | 6 (6.5) | 68 (73.1) | 20<br>(16.4) | 11 (9.0) | 91 (74.6) | .028 | .1282 |
| congenital heart abnormalities (n=329) | 64<br>(19.3) | 14 (4.2) | 253 (76.4) | 22<br>(19.1) | 1 (0.9) | 92 (80.0) | 22<br>(23.4) | 4 (4.3) | 68 (72.3) | 20<br>(19.4) | 9 (7.4) | 93 (76.2) | .104 |  |
| caesarean delivery (n=329) | 66<br>(20.1) | 17 (5.2) | 246 (74.8) | 19<br>(16.4) | 0 (0.0) | 97 (83.6) | 22<br>(23.7) | 5 (5.4) | 66 (71.0) | 25<br>(20.8) | 12 (10.0) | 83 (69.2) | .005 | .1497 |
| macrosomia (n=329) | 56<br>(17.0) | 28 (8.5) | 245 (74.5) | 21<br>(18.1) | 1 (0.9) | 94 (81.0) | 17<br>(18.5) | 4 (4.4) | 71 (77.2) | 18<br>(14.9) | 23 (19.0) | 80 (66.1) | <.001 | .2060 |
| labour dystocia (n=329) | 64<br>(19.5) | 24 (7.3) | 241 (73.3) | 19<br>(16.5) | 1 (0.9) | 95 (82.6) | 20<br>(21.5) | 3 (3.2) | 70 (75.3) | 25<br>(20.7) | 20 (16.5) | 76 (62.8) | <.001 | .2018 |
| stillbirth (n=327) | 68<br>(20.8) | 20 (6.1) | 239 (73.1) | 25<br>(22.1) | 1 (0.9) | 87 (77.0) | 18<br>(19.6) | 5 (5.4) | 69 (75.0) | 25<br>(20.5) | 14 (11.5) | 83 (68.0) | .019 | .1339 |

| Influence of maternal preconception | Total |  |  | GP |  |  | Midwives |  |  | Naturopaths |  |  | P** | Cramer |
| --- | --- | --- | --- | --- | --- | --- | --- | --- | --- | --- | --- | --- | --- | --- |
| medically induced labour (n=332) | 71<br>(21.4) | 20 (6.0) | 241 (72.6) | 26<br>(22.4) | 0 (0.0) | 90<br>(77.6) | 21<br>(22.3) | 4 (4.3) | 69<br>(73.4) | 24<br>(19.7) | 16<br>(13.1) | 82 (67.2) | .001 | .1683 |
| cryptorchidism (n=330) | 66<br>(20.0) | 29 (8.8) | 235 (71.2) | 21<br>(18.3) | 0 (0.0) | 94<br>(81.7) | 23<br>(24.7) | 7 (7.5) | 63<br>(67.7) | 22<br>(18.0) | 22<br>(18.0) | 78 (64.0) | <.001 | .2002 |
| precipitous labour (n=330) | 77<br>(23.3) | 24 (7.3) | 229 (69.4) | 25<br>(21.6) | 0 (0.0) | 91<br>(78.5) | 23<br>(24.7) | 3 (3.2) | 67<br>(72.0) | 29<br>(24.0) | 21<br>(17.4) | 71 (58.7) | <.001 | .2173 |
| shoulder dystocia (n=331) | 79<br>(23.9) | 26 (7.9) | 226 (68.3) | 26<br>(22.4) | 0 (0.0) | 90<br>(77.6) | 17<br>(18.3) | 4 (4.3) | 72<br>(77.4) | 36<br>(29.5) | 22<br>(18.0) | 64 (52.5) | <.001 | .2355 |
| ...on child outcomes |  |  |  |  |  |  |  |  |  |  |  |  |  |  |
| neurocognitive development (n=329) | 53<br>(16.1) | 8 (2.4) | 268 (81.5) | 24<br>(21.1) | 1 (0.9) | 89<br>(78.1) | 17<br>(18.1) | 4 (4.3) | 73<br>(77.7) | 12 (9.9) | 3 (2.5) | 106 (87.6) | .086 |  |
| emotional/behavioural problems (n=330) | 62<br>(18.8) | 9 (2.7) | 259 (78.5) | 23<br>(19.8) | 0 (0.0) | 93<br>(80.2) | 18<br>(19.2) | 5 (5.3) | 71<br>(75.5) | 21<br>(17.5) | 4 (3.3) | 95 (79.2) | .201 |  |
| developmental delay (n=331) | 65<br>(19.6) | 9 (2.7) | 257 (77.6) | 24<br>(20.9) | 0 (0.0) | 91<br>(79.1) | 20<br>(21.3) | 5 (5.3) | 69<br>(73.4) | 21<br>(17.2) | 4 (3.3) | 97 (79.5) | .165 |  |
| asthma (n=330) | 69<br>(20.9) | 9 (2.7) | 252 (76.4) | 25<br>(21.6) | 1 (0.9) | 90<br>(77.6) | 23<br>(24.5) | 5 (5.3) | 66<br>(70.2) | 21<br>(17.5) | 3 (2.5) | 96 (80.0) | .217 |  |
| attention deficit-hyperactivity disorder (n=329) | 68<br>(20.7) | 10 (3.0) | 251 (76.3) | 26<br>(22.6) | 1 (0.9) | 88<br>(76.5) | 19<br>(20.4) | 4 (4.3) | 70<br>(75.3) | 23<br>(19.0) | 5 (4.1) | 93 (76.9) | .535 |  |
| overweight (n=327) | 72<br>(22.0) | 7 (2.1) | 248 (75.8) | 24<br>(21.4) | 0 (0.0) | 88<br>(78.6) | 27<br>(29.0) | 3 (3.2) | 63<br>(67.7) | 21<br>(17.2) | 4 (3.3) | 97 (79.5) | .086 |  |
| gross motor function (n=327) | 76<br>(23.2) | 9 (2.8) | 242 (74.0) | 25<br>(21.6) | 0 (0.0) | 91<br>(78.5) | 27<br>(30.0) | 4 (4.4) | 59<br>(65.6) | 24<br>(19.8) | 5 (4.1) | 92 (76.0) | .068 |  |
| cerebral palsy (n=329) | 74<br>(22.5) | 14 (4.3) | 241 (73.3) | 20<br>(17.7) | 0 (0.0) | 93<br>(82.3) | 23<br>(24.5) | 4 (4.3) | 67<br>(71.3) | 31<br>(25.4) | 10 (8.2) | 81 (66.4) | .011 | .1413 |
| autism spectrum (n=331) | 77<br>(23.3) | 13 (3.9) | 241 (72.8) | 26<br>(22.4) | 1 (0.9) | 89<br>(76.7) | 22<br>(23.4) | 6 (6.4) | 66<br>(70.2) | 29<br>(24.0) | 6 (5.0) | 86 (71.1) | .288 |  |
| leukaemia (n=327) | 86<br>(26.3) | 22 (6.7) | 219 (67.0) | 28<br>(24.4) | 3 (2.6) | 84<br>(73.0) | 22<br>(23.9) | 7 (7.6) | 63<br>(68.5) | 36<br>(30.0) | 12<br>(10.0) | 72 (60.0) | .114 |  |

\*Unlikely = Extremely unlikely and moderately unlikely; Unsure = Slightly unlikely, slightly likely and unsure; Likely = Moderately likely and extremely likely
\*\*The Cramer's V and P values reflect statistical tests examining differences in health literacy between health professions

Infant health outcomes that participants most commonly accurately identified as ‘likely’ to be influenced by maternal preconception exposures were low birth weight (83.7%), small for gestational age (81.1%) and neural tube defects (80.6%) while the outcome identified most as ‘unlikely’ was shoulder dystocia (23.9%) and precipitous labour (23.3%). The infant outcomes for which participants were most ‘uncertain’ were cryptorchidism (8.8%) and macrosomia (8.5%).

Participants most commonly identified neurocognitive development (81.5%) and emotional or behavioural problems (78.5%) as outcomes affecting the child due to maternal preconception exposures. Leukaemia was the outcome of maternal preconception exposures that the most participants identified incorrectly (‘unlikely’) (26.3%) or were unsure (6.7%) about.

Differences in accuracy by profession was seen in 75% of infant outcomes of which five outcomes had a moderate association: shoulder dystocia (V=.2355), precipitous labour (V=.2173), macrosomia (V=.2060), labour dystocia (V=.2018) and cryptorchidism (V=.2018). GPs were the profession with the highest proportion of participants that identified ‘likely’ for each of these outcomes (77.6% - 82.6%), although midwives had a similar proportion of respondents choosing ‘likely’ for shoulder dystocia (GPs: 77.6%, midwives: 77.4%) while naturopaths were consistently lowest for all infant outcomes with a moderate effect size (52.5% - 66.1%). Accurate perceptions of the likely impacts of maternal preconceptions exposures on one child health outcome, cerebral palsy, was weakly associated with health profession category (V=.1413), and similar to the infant health outcomes, GPs most frequently identified maternal preconception exposures as likely to influence this outcome (82.3%) while naturopaths had the lowest proportion (66.4%). No differences in responses between health professions were identified for pregnancy and postnatal outcomes.

### Knowledge the relationship between paternal preconception risks and offspring outcomes

Table 6 presents participant responses regarding paternal preconception exposures and outcomes. The risk factor identified by most participants as impacting offspring outcomes was paternal chemical exposure in the preconception period (likely: 82.7%), while paternal preconception BMI received the highest rate of ‘unlikely’ (18.8%) responses.

**Table 6:**
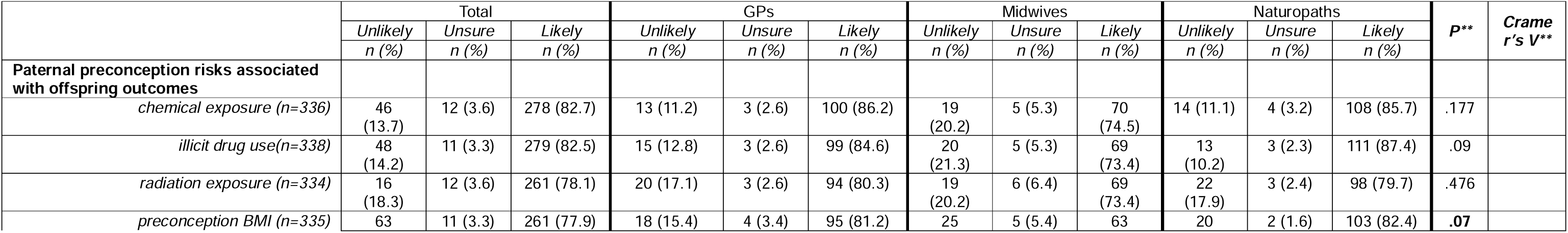
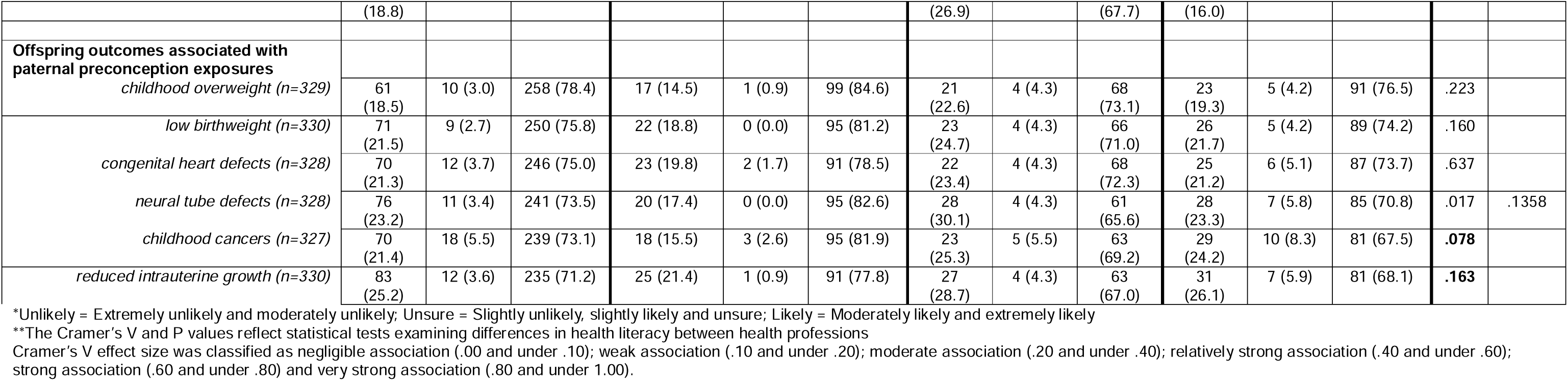
Knowledge of influence of paternal preconception risks and outcomes associated with paternal preconception exposures.

The influence of paternal preconception risks on infant or child health outcomes found the highest level of accuracy identified paternal preconception risks as impacting on childhood overweight (78.4%) while the most ‘unlikely’ responses were for reduced intrauterine growth (25.2%) and the highest uncertainty was for childhood cancers (5.5%).

A weak association with health profession group was found for perceived likelihood of paternal preconception exposures contributing to neural tube defects (V=.1358) but no other outcomes. The proportion of respondents identifying neural tube defects as ‘likely’ associated with paternal preconception exposures were highest for GPs (82.6%) and lowest for midwives (65.6%).

## Discussion

This study offers critical insights into the preconception information-seeking behaviours and health literacy of three groups of primary care health professionals-GPs, midwives, and naturopaths - who deliver preconception care in clinical settings across Australia.

### Preconception knowledge gaps among primary care professionals

Overall, the findings of this study suggest that primary care clinicians are generally aware of maternal and paternal preconception risks, exposures, and associated maternal, infant and child health outcomes. Rates of participant responses aligned with the evidence commonly ranged between 60% and 90%, indicating an overall adequacy in preconception health knowledge among participants. Variability between professions was also identified, the most prominent seen in participants’ understanding of infant outcomes associated with maternal preconception exposures. It is worth noting, however, that the preconception health exposures and outcomes now recognised in the literature is broad and continuing to grow as research interest in this topic area increases.^1^ This evolution in the evidence-base presents challenges not faced among health issues with more focused, established and stable content (e.g., cervical screening).^26^ These ongoing changes can further exacerbate the current gaps in knowledge identified through this study, and may require educators and policymakers to take lessons from topics that are more accurately understood by health professions, to create more universality to their preconception health literacy. The highest level of accurate health knowledge, for example, was related to the association between maternal preconception alcohol intake and offspring outcomes. Participant understanding of alcohol use may be influenced by the health promotion campaigns targeting maternal alcohol consumption during pregnancy in Australia, despite the challenges faced in implementing these campaigns.^27^ In contrast, a lower proportion of participants identified folic acid in the preconception period as impacting offspring outcomes in the face of targeted public health and health promotion interventions to educate the community and health professionals in this regard.^28^ Clinical practice guidelines, based on high-quality evidence, in particular, recommend folic acid for the prevention of neural tube defects.^29^ Yet 1 in 5 participants did not consider fetal development issues resulting in neural tube defects ‘likely’ to be linked to maternal preconception outcomes. The results also showed that more than one quarter of participants did not identify that folic acid intake was likely to impact offspring outcomes. Ultimately these specific examples, taken in the context of the overall results, highlight the participants’ inconsistent understanding of both preconception exposures and outcomes and need to be further investigated to inform health practitioner preconception care skills development.

### Clinical preconception information and education sources require quality standards

In our study, clinicians indicated they mainly obtained their preconception health knowledge through continuing professional education provided by clinicians or researchers. They predominantly accessed professional journals and clinical textbooks when seeking information on preconception health. These findings align with previous qualitative research involving GPs, midwives and naturopaths in which participants described gaining the majority of their knowledge regarding preconception health and care through professional development courses and seminars.^30^ Interestingly, clinical practice guidelines were not as popular a source of preconception health information for participants, despite this being the preferred format for policy and evidence-based practice. A recent examination of preconception care clinical practice guidelines has also identified weaknesses in existing guidelines including not drawing on the most robust evidence, and omitting important topics such as paternal preconception exposures and outcomes.^29^ Participants did, however, describe colleagues, professional training, and clinical experience as important avenues for acquiring knowledge on preconception health and care.^30^ However, the quality of the information provided through these more tacit sources is not clear and needs research attention. Certainly, any training or resources being developed should be codesigned with clinicians to ensure they meet their knowledge gaps and practice needs. Preconception health care could be developed for undergraduate medical and health sciences clinical curricula to increase preconception health literacy and clinical awareness for primary care practitioners. Such training should provide consistent content for all primary care clinicians – including and beyond those sampled for this study - while also permitting sufficient flexibility to adapt to the unique learning needs and clinical scope of different professions.

### Clinicians need more preconception health resources to support care

Despite our participants’ identified preconception health knowledge gaps and challenges accessing preconception health information, it is also interesting to note that participants consider patient literacy to be a greater barrier to preconception care than clinician literacy. Given the poor awareness and understanding about the importance and impacts of preconception health in the community,^31^ this view is likely justified. For this reason, efforts to strengthen the primary care capability to deliver preconception care needs to be coupled with broader health promotion interventions to educate, inform and motivate the general population.^32^ In line with previous research^33^ and our participants suggestions, such campaigns can be integrated with primary care by ensuring clinicians have greater access to patient brochures and handouts. In an Australian qualitative study of the same three primary care professions included in our study (i.e., GPs, midwives and naturopaths), participants reported using handouts to provide their patients with preconception health information.^34^ It is unclear where these previous study participants sourced their patient handouts as there are no formal government or professional preconception health patient education materials available in Australia. State agencies have developed some public-facing websites however the longest standing site established by the Victorian Assisted Reproductive Technology Agency was defunded in 2024^35^ - albeit still live at the time of the survey - while a new website developed by Health and Wellbeing Queensland in 2026^36^ was not yet active.

### Limitations and strengths

This study has several limitations. Methodological limitations include being an online voluntary, self-reported survey and which exposes the study to both selection and recall bias. Participants may have a special interest in the topic compared to other health professionals, which could potentially bias results. The participants represented three primary care health professions: GPs, midwives, and naturopaths. The findings may not reflect other health professions’ preconception health literacy or associated information-seeking behaviours. These limitations may impact the generalisation of the findings. Despite these limitations, this study offers a valuable examination of GPs’, midwives’, and naturopaths’ preconception information-seeking behaviour and their knowledge of maternal and paternal preconception risk factors, pregnancy outcomes, and health outcomes for both mothers and offspring within Australian primary care settings.

## Conclusions

Overall while there may be adequate preconception health literacy among these primary care practitioners the necessary knowledge is far from universally accurate across exposure, outcomes and between health professions. The level of health literacy of some key preconception health behaviours, such as maternal folic acid, is concerning. Primary care clinicians require more evidence-based resources and education related to preconception health to effectively provide care. They need greater access to, and awareness of, preconception care clinical practice guidelines that address both women’s and men’s health risks and exposures, or to other codesigned materials that suit their knowledge and practice needs. Preconception care clinical practice guidelines should be tailored to the scope of practice for each respective profession – noting that no one profession demonstrated a high level of accuracy across all topics. Patient education resources also need to be developed for primary care and should be codesigned with the community and primary care clinicians to ensure they meet the needs of both. Finally, clinicians need access to comprehensive preconception care education that is suited to the diverse scope of practice and experience seen among different primary care professions.

## Data Availability

All data produced in the present study are available upon reasonable request to the authors

## List of abbreviations

ACM: Australian College of Midwives
CMA: Complementary Medicines Association
GP: General practitioner
NHAA: Naturopaths and Herbalists Association of Australia
RACGP: Royal Australian College of General Practitioners

## Declarations

### Ethics approval and consent to participate

The research was approved by the University of Technology Sydney Human Research Ethics Committee (ETH22-7083).

### Consent for publication

Not applicable

### Availability of data materials

The datasets used and/or analysed during the current study are available from the corresponding author upon reasonable request.

### Author’ contributions

The authors confirm their contribution to the paper: study conception and design: CC, AS, DS AND EM; data collection: CC, AS; analysis and interpretation of results: CC, AS; draft manuscript preparation: CC, AS, DS AND EM. All authors reviewed the results and approved the final version of the manuscript.

## Acknowledgements

The authors thank member associations RACGP, RACGP Specific Interest group Antenatal and Postnatal Care, ACM, NHAA and CMA representing GPs, midwives, and naturopaths for supporting this research by promoting the study to their members for participation. The authors also thank each health professional who gave their time to participate in the study. The authors thank Endeavour College of Natural Health for supporting the research study by awarding a project grant.

